# Predictive performance and clinical application of COV50, a urinary proteomic biomarker in early COVID-19 infection: a cohort study

**DOI:** 10.1101/2022.01.20.22269599

**Authors:** Jan A. Staessen, Ralph Wendt, Yu-Ling Yu, Sven Kalbitz, Lutgarde Thijs, Justyna Siwy, Julia Raad, Jochen Metzger, Barbara Neuhaus, Armin Papkalla, Heiko von der Leyen, Alexandre Mebazaa, Emmanuel Dudoignon, Goce Spasovski, Mimoza Milenkova, Aleksandra Canevska-Taneska, Mina Psichogiou, Marek W. Rajzer, Lukasz Fulawka, Magdalena Dzitkowska-Zabielska, Guenter Weiss, Torsten Feldt, Miriam Stegemann, Johan Normark, Alexander Zoufaly, Stefan Schmiedel, Michael Seilmaier, Benedikt Rumpf, Mirosław Banasik, Magdalena Krajewska, Lorenzo Catanese, Harald Rupprecht, Beata Czerwienska, Björn Peters, Åsa Nilsson, Katja Rothfuss, Christoph Lübbert, Harald Mischak, Joachim Beige, the CRIT-Cov-U investigators

**Affiliations:** Non-Profit Research Institute Alliance for the Promotion of Preventive Medicine, Mechelen, Belgium; Biomedical Sciences Group, Faculty of Medicine, University of Leuven, Leuven, Belgium; Department of Infectious Diseases/Tropical Medicine, Nephrology/KfH Renal Unit and Rheumatology, St. Georg Hospital Leipzig, Germany; Research Unit Environment and Health, Department of Public Health; Research Unit Hypertension and Cardiovascular Epidemiology, Department of Cardiovascular Diseases, University of Leuven, Leuven, Belgium; Mosaiques-Diagnostics GmbH, Hannover, Germany; Center for Clinical Trials - CTS, Medizinische Hochschule, Hannover, Germany; Department of Anaesthesiology and Intensive Care, Hospital Saint Louis-Lariboisière, Paris, France; Cyril and Methodius University, Skopje, Republic of North Macedonia; First Department of Internal Medicine, Laiko General Hospital, National and Kapodistrian University of Athens Medical School, Athens, Greece; First Department of Cardiology, Interventional Electrocardiology and Arterial Hypertension, Jagiellonian University Medical College, Kraków, Poland; Molecular Pathology Centre Cellgen, Wroclaw, Poland; Faculty of Physical Education, Gdansk University of Physical Education and Sport; Center of Translational Medicine, Medical University of Gdańsk, Poland; Department of Infectious Diseases and Respiratory Medicine, Charité – Universitätsmedizin Berlin, corporate member of Freie Universität Berlin and Humboldt-Universität zu Berlin, Germany; Department of Internal Medicine II, Medical University Innsbruck, Innsbruck, Austria; Department of Gastroenterology, Hepatology and Infectious Diseases, Medical Faculty of Heinrich Heine University, Düsseldorf, Germany; Wallenberg Centre for Molecular Medicine, Department of Clinical Microbiology, Umeå University, Umeå, Sweden; Department of Medicine IV, Clinic Favoriten and Faculty of Medicine, Sigmund Freud University, Vienna, Austria; Nephrology and Dialysis, Internal Medicine III, Medical University of Vienna, Vienna, Austria; Department of Nephrology and Transplantation Medicine, Wroclaw Medical University, Wroclaw, Poland; University of Silesia, Katowice, Poland; Department of Nephrology, Skaraborg Hospital, Skövde and Department of Molecular and Clinical Medicine, Institute of Medicine, The Sahlgrenska Academy at University of Gothenburg, Sweden; Research and Development Centre, Skaraborg Hospital, Skövde, Sweden; Department of Nephrology, Angiology and Rheumatology, Hospital Bayreuth; Margarete-Fischer-Bosch Institute for Clinical Pharmacology and Division of Infectious Diseases, Robert Bosch Hospital, Stuttgart, Germany; Medical Department I and Bernhard-Nocht-Clinic for Tropical Medicine, University Medical Center Hamburg Eppendorf, Hamburg, Germany; Department of Haematology, Oncology, Immunology, Palliative Care, Infectious Disease and Tropical Medicine, München Klinik Schwabing, München, Germany; Division of Infectious Diseases and Tropical Medicine, Leipzig University Medical Center, Leipzig, Germany; Institute of Cardiovascular and Medical Sciences, Glasgow, United Kingdom; Martin-Luther-University Halle-Wittenberg, Halle an der Saale, Germany

**Keywords:** COVID-19, disease severity, risk score, SARS-CoV-2, urinary proteomics, World Health Organization scale

## Abstract

**Background:** The SARS-CoV-2 pandemic remains a worldwide challenge. The CRIT-Cov-U pilot study generated a urinary proteomic biomarker consisting of 50 peptides (COV50), which predicted death and disease progression. Following the interim analysis demanded by the German government, the full dataset was analysed to consolidate findings and propose clinical applications.

**Methods:** In eight European countries, 1012 adults with PCR-confirmed COVID-19 were followed up for death and progression along the 8-point WHO scale. Capillary electrophoresis coupled with mass spectrometry was used for urinary proteomic profiling. Statistical methods included logistic regression, receiver operating curve analysis with comparison of the area under curve (AUC) between nested models. Hospitalisation costs were derived from the care facility corresponding with the Markov chain probability of reaching WHO scores ranging from 3 to 8 and flat-rate hospitalistion costs standardised across countries.

**Findings:** The entry WHO scores were 1-3, 4-5 and 6 in 445 (44·0%), 529 (52·3%), and 38 (3·8%) patients, of whom 119 died and 271 progressed. The standardised odds ratios associated with COV50 for death were 2·44 (95% CI, 2·05-2·92) unadjusted and 1·67 (1·34-2·07) if adjusted for sex, age, body mass index, comorbidities and baseline WHO score, and 1·79 (1·60-2·01) and 1·63 (1·40-1·90), respectively, for disease progression (p<0·0001 for all). The predictive accuracy of optimised COV50 thresholds were 74·4% (95% CI, 71·6-77·1) for mortality (threshold 0·47) and 67·4% (64·1-70·3) for disease progression (threshold 0·04). On top of covariables and the baseline WHO score, these thresholds improved AUCs from 0·835 to 0·853 (p=0·0331) and from 0·697 to 0·730 (p=0·0008) for death and progression, respectively. Of 196 ambulatory patients, 194 (99·0%) did not reach the 0·04 threshold. Earlier intervention guided by high-risk COV50 levels should reduce hospital days with cost reductions expressed per 1000 patient-days ranging from M€ 1·208 (95% percentile interval, 1·035-1·406) at low risk (COV50 <0·04) to M€ 4·503 (4·107-4·864) at high risk (COV50 ≥0·04 and age ≥65 years).

**Interpretation:** The urinary proteomic COV50 marker is accurate in predicting adverse COVID-19 outcomes. Even in mild-to-moderate PCR-confirmed infections (WHO scores 1-5), the 0·04 threshold justifies earlier drug treatment, thereby reducing hospitalisation days and costs.

**Funding:** German Federal Ministry of Health acting upon a decree from the German Federal Parliament.

## Introduction

The SARS-CoV-2 pandemic remains a challenge for health care worldwide. Globally, during the week of 15-21 November 2021, nearly 3·6 million new cases and over 51,000 deaths were reported, reflecting 6% increases in both metrics compared to the preceding week.^1^ While the pandemic is stretching health care resources over what is sustainable in the long run, the world is now bracing for the Omicron variant with higher transmissibility and potentially more resistance against the immunological response to vaccines or previous infection.^2,3^ Until now COVID-19 patients were stratified for risk based on age, obesity, comorbidities and disease-severity scales.

An extensive literature search (appendix 2) did not reveal any predictive biomarker allowing to select high-risk COVID-19 patients for early treatment. Urinary proteomic profiling (UPP) generates classifiers representative for pathogenic molecular mechanisms, which in case of SARS-CoV-2 infection are generally independent of mutating virus strains and which may inform treatment, in particular with pharmacological agents not specifically directed against the variable S-protein domains of evolving SARS-CoV2 strains. Following a insistent request from the German government, an interim analysis of the Prospective Validation of a Proteomic Urine Test for Early and Accurate Prognosis of the Critical Complications in Patients with SARS-CoV-2 Infection Study (CRIT-Cov-U) described the discovery, replication and internal and external validation of COV50. This novel UPP biomarker consists of 50 deregulated urinary peptides (appendix 1 p 8) and predicts death and progression across the COVID-19 WHO stages over beyond risk factors and comorbidities.^4^ The objectives of the current study were to consolidate the interim findings in the full study sample and to propose potential applications of the COV50 marker in clinical practice and trial design.

## Methods

CRIT-Cov-U project complies with the Helsinki declaration. The Ethics Committee of the German-Saxonian Board of Physicians, Dresden, Germany (number, EK-BR-88/20.1) and the Institutional Review Boards of the recruiting sites provided ethical clearance. The protocol is deposited at the German Register for Clinical Studies (www.drks.de; number DRKS00022495), which is linked to the WHO International Clinical Trial Registry Platform (www.who.int/clinical-trials-registry-platform).

CRIT-Cov-U was a prospective multicentre cohort study.^4^ Eligible patients were non-anuric adults (≥18 years), capable to give written informed consent, with PCR-confirmed SARS-CoV-2 infection diagnosed in ambulatory care or on the first hospitalisation day. All patients meeting the eligibility criteria were enrolled without any exclusion in two phases: 228 patients were recruited from 25 March 2020 until 18 November 2020 and were included in the interim report;^4^ recruitment of 784 patients continued from 30 April 2020 until 14 April 2021, so that the full study cohort comprised 1012 individuals. Five centres participated in the initial enrolment of patients and an additional 12 in the continued recruitment. Two sites were located in Austria (65 patients enrolled), one in France (49), seven in Germany (458), one in Greece (30), one in North Macedonia (137), four in Poland (149), one in Spain (23), and two in Sweden (101).

All patients were followed up until recovery, hospital discharge or death. On days 0-3, 4-7 and 10-21 after diagnosis surviving patients were staged according to the 8-point WHO Clinical Progression Scale.^5^ Electronic case report forms (MARVIN EDC, XClinical GmbH, Munich, Germany) were used for data compilation. For UPP, 8 ml-urine samples were collected in borated test tubes (ExactoBac-U®, Sarstedt, Nümbrecht, Germany) and kept at -20 °C until assayed. The methods for capillary electrophoresis coupled with mass spectrometry, peptide sequencing, and for evaluation, calibration and quality control of the mass spectrometric data are described in detail in appendix 1 (p 2-5).

The initial sample size calculations informed by a proof-of-concept study^6^ required 212 patients with critical COVID-19 (WHO stage ≥6) to be contrasted with 271 patients with mild symptoms (WHO stage <4) to identify and validate a UPP biomarker having 75% sensitivity and 80% specificity. Given the 33% progression rate from mild to severe disease in the pilot study^6^ and accounting for 15% of missing data, the sample size for the full study was set at 645 patients. Based on the interim study,^4^in which the mortality rate was 10·1% (23/228), the sample size for the full study was revised to 1000 patients.

For database management and statistical analysis, SAS software version 9·4 was used. Significance was a 2-tailed significance of 0·05 or less. Means and proportions were compared using the large-sample z test or ANOVA and Fisher’s exact test, respectively. The predefined endpoints were mortality and progression across the 8-point WHO scale of disease severity. The 95% confidence intervals of rates were computed as R ± 1.96 × √(R/T), where R and T are the rate and the denominator used to calculate the rate. The risk of incident endpoints was related to the baseline COV50 level by logistic regression, unadjusted or adjusted for sex, age, the entry WHO scale and comorbidities including hypertension, heart failure, diabetes, and cancer. The differences in the COV50 odds ratios between initial and continued recruitment were tested by introduction of the interaction between study phase and baseline COV50 in the logistic models. Performance of COV50 in risk stratification was assessed by the area (AUC) under the receiver operating curve (ROC) and the Delong approach to compare the AUCs between nested models. The COV50 thresholds optimised by the Youden index were 0·47 for mortality and 0·04 for worsening WHO score.^4^

A Markov chain analysis was conducted using SAS IML and bootstrapped 1000 times to generate the transition probabilities from the entry to follow-up WHO scores (appendix 1 p 17).^7^ The transition probabilities were computed for various risk strata defined by the entry COV50 level and age (<65 *vs* ≥65 years). The transition probabilities allowed extrapolating the number of patients reaching follow-up WHO scores of 3-4, 5 and 6-8, and therefore requiring regular, intermediate or intensive care. Given the simulated number of patients, hospitalisation costs were computed based on the median number of hospital days per care facility, as observed in the CRIT-Cov-U cohort, and a flat care-facility specific costing rate, which was harmonised across clinical sites based on each country’s 2020 gross domestic product^8^ with the diagnosis-related daily costs applicable in Germany as reference: €540, €1590, and €1770 for regular, intermediate and intensive care, respectively. Hospitalisation costs were expressed in million Euro (M€) for 1000 patients in each risk stratum.

## Results

The 1012 participants making up the full dataset were on average 62·3 years old, included 447 (44·2%) women, 557 (55·0%) patients with hypertension, 154 (15·2%) with heart failure, 257 (25·4%) with diabetes, and 106 (10·5%) patients with cancer (appendix 1 p 18). The WHO score at enrolment (table 1) was 1-3 in 445 (44·0%) patients, 4-5 in 529 (52·3%), and 6 in 38 (3·8%). The median (interquartile range) COV50 level at baseline was 0·23 (−1.27 to 0·80; appendix 1 p 19). Compared with the initially enrolled patients, those recruited later scored lower on the WHO scale (table 1; p<0·0001), included more patients with a history of cancer (5·7 *vs* 11·9%; p=0·0117), but fewer participants on inhibitors of the renin-angiotensin system (53·5% *vs* 38·9%; p<0·0001). Otherwise, patients recruited initially and later had similar characteristics (table 1), in particular a comparable entry level of COV50 (−0·19 *vs* -0·24; p=0·5918). In the whole study population (appendix 1 p 11), the proportion of women and the mean values of diastolic blood pressure decreased across increasing fourths of the COV50 distribution, whereas age, heart rate and the rates of hypertension, heart failure, diabetes mellitus and cancer increased.

**Table 1:** Baseline characteristics.

| Characteristic | Recruitment phase |  |  | Full study |
| --- | --- | --- | --- | --- |
|  | Initial | Continued | p value |  |
| Number in cohort | 228 | 784 |  | 1012 |
| Main study variables |  |  |  |  |
| WHO score |  |  |  |  |
| 1-3 | 90 (39.5) | 355 (45.3) |  | 445 (44.0) |
| 4-5 | 107 (46.9) | 422 (53.8) | <0.0001 | 529 (52.3) |
| 6 | 31 (13.6) | 7 (0.9) |  | 38 (3.8) |
| COV50 level | -0.19 (1.52) | -0.24 (1.36) | 0.5918 | -0.23 (1.40) |
| Number with characteristic (%) |  |  |  |  |
| White ethnicity | 205 (89.9) | 685 (87.4) | 0.2999 | 890 (87.9) |
| Women | 94 (41.2) | 353 (45.0) | 0.3095 | 447 (44.2) |
| Hypertension | 137 (60.1) | 420 (53.6) | 0.0817 | 557 (55.0) |
| Heart failure | 30 (13.6) | 124 (15.8) | 0.3253 | 154 (15.2) |
| Body mass index $\geq 30$ kg/m<sup>2</sup> | 59 (29.8) | 192 (24.5) | 0.6694 | 251 (24.8) |
| Diabetes mellitus | 65 (28.5) | 192 (24.5) | 0.2198 | 257 (25.4) |
| Cancer | 13 (5.7) | 93 (11.9) | 0.0117 | 106 (10.5) |
| Use of RAS blockers, | 122 (53.5) | 305 (38.9) | <0.0001 | 427 (42.2) |
| Mean (SD) of characteristic |  |  |  |  |
| Age | 63.1 (17.1) | 62.1 (18.0) | 0.4556 | 62.3 (17.8) |
| Systolic blood pressure, mm Hg | 129.8 (23.2) | 127.7 (19.0) | 0.1633 | 128.2 (20.1) |
| Diastolic blood pressure, mm Hg | 75.9 (13.5) | 76.2 (11.7) | 0.7436 | 76.2 (12.2) |
| Heart rate, beats per minute | 83.4 (15.1) | 81.9 (15.6) | 0.2074 | 82.2 (15.5) |
| Body mass index, kg/m<sup>2</sup> | 28.0 (5.4) | 27.5 (5.2) | 0.2252 | 27.6 (5.2) |

RAS blockers indicate blocker of the renin-angiotensin system, including angiotensin-converting enzyme inhibitors and angiotensin-receptor blockers. Systolic and diastolic blood pressure and heart rate were missing in 2 initially recruited participants and 29 participants recruited later. The p value refers to the differences in the study participants' characteristics between initial recruitment (25 March 2020 – 18 November 2020) and continued recruitment (30 April 2020 – 14 April 2021). RAS=renin-angiotensin system.

No patient was lossed to follow-up. Median (5th-95th percentile interval) follow-up was 10 days (1-34) for mortality (119 deaths) and 10 days (2-26) for worsening WHO score (271 cases). The baseline COV50 distribution shifted upward (p<0·0001), when plotted against the worst WHO score attained during follow-up (appendix 1 p 20). In the whole study population (table 2), the relative risk of death expressed per 1-SD increment in COV50 at entry was 2·44 (95% confidence interval, 2·05-2·92; p<0·0001) unadjusted and 1·67 (1·34-2·07) when fully adjusted for sex, age, body mass index, the presence of comorbidities, and the baseline WHO score; for progression in the WHO score (table 3), the corresponding odds ratios were 1·79 (1·60-2·01) unadjusted and 1·63 (1·10-1·90) fully adjusted. In analyses dichotomised by study phase, the risk associated with COV50 was similar for both endpoints, irrespective of adjustment (table 2). The unadjusted odds ratios for initial compared with continued recruitment were 2·45 (1·69-3·54) *vs* 2·47 (2·02-3·03) for mortality (p=0·9605) and 1·95 (1·52-2·51) *vs* 1·77 (1·56-1·73) for worsening WHO score (p=0·5150). The corresponding fully adjusted estimates (table 2) were 2·27 (1·34-3·83) *vs* 1·55 (1·21-1·98) for mortality (p=0·9380) and 1·95 (1·52-2·51) *vs* 1·77 (1·56-1·73) for worsening WHO score (p=0·1645).

**Table 2:** Odds ratios relating outcome to COV50 by recruitment phase.

| Outcome | Initial |  | Continued |  | Full |  |
| --- | --- | --- | --- | --- | --- | --- |
|  | OR (95% CI) | p value | OR (95%CI) | p value | OR (95% CI) | p value |
| **Mortality** |  |  |  |  |  |  |
| N° deaths/at risk (%) | 25/228 (11·0) |  | 94/784 (12·0) |  | 119/1012 (11·8) |  |
| Unadjusted | 2·45 (1·69-3·54) | <0·0001 | 2·47 (2·02-3·03) | <0·0001 | 2·44 (2·05-2·92) | <0·0001 |
| Adjusted |  |  |  |  |  |  |
| Sex and age | 2·30 (1·57-3·37) | <0·0001 | 1·88 (1·50-2·35) | <0·0001 | 2·04 (1·68-2·47) | <0·0001 |
| + baseline WHO score | 2·18 (1·30-3·64) | 0·0030 | 1·54 (1·21-1·96) | 0·0005 | 1·65 (1·34-2·05) | <0·0001 |
| + BMI and comorbidities | 2·27 (1·34-3·83) | 0·0023 | 1·55 (1·21-1·98) | 0·0005 | 1·67 (1·34-2·07) | <0·0001 |
| **Progressing WHO score** |  |  |  |  |  |  |
| N° events/at risk (%) | 50/228 (21·9) |  | 221/784 (28·3) |  | 271/1012 (26·8) |  |
| Unadjusted | 1·95 (1·52-2·51) | <0·0001 | 1·77 (1·56-2·02) | <0·0001 | 1·79 (1·60-2·01) | <0·0001 |
| Adjusted |  |  |  |  |  |  |
| Sex and age | 1·81 (1·38-2·35) | <0·0001 | 1·50 (1·29-1·73) | <0·0001 | 1·56 (1·38-1·77) | <0·0001 |
| + baseline WHO score | 2·32 (1·56-3·46) | <0·0001 | 1·52 (1·29-1·79) | <0·0001 | 1·65 (1·42-1·92) | <0·0001 |
| + BMI and comorbidities | 2·32 (1·55-3·48) | <0·0001 | 1·51 (1·27-1·78) | <0·0001 | 1·63 (1·40-1·90) | <0·0001 |

Odds ratios given with 95% confidence interval express the risk for 1-SD increment in COV50. Initial recruitment lasted from 25 March 2020 until 18 November 2020 and continued recruitment from 30 April 2020 until 14 April 2021. Comorbidities include hypertension, heart failure, diabetes and cancer. BMI=body mass index.

**Table 3:** Discriminative performance of COV50 by recruitment phase.

| Outcome | Initial | Continued | Full |
| --- | --- | --- | --- |
| **Mortality** |  |  |  |
| N° deaths/at risk (%) | 25/228 (11·0) | 94/784 (12·0) | 119/1012 (11·8) |
| Continuously distributed COV50 |  |  |  |
| AUC (95% confidence interval) | 0·83 (0·77-0·90) | 0·83 (0·77-0·90) | 0·81 (0·77-0·85) |
| Categorised COV50 |  |  |  |
| Youden cut-off threshold | 0·47 | 0·47 | 0·47 |
| Sensitivity (95% confidence interval) | 88·0 (68·8-97·4) | 71·3 (61·0-80·1) | 74·8 (66·0-82·3) |
| Specificity (95% confidence interval) | 75·4 (68·9-81·1) | 75·5 (72·2-78·6) | 74·4 (71·4-77·2) |
| PLR (95% confidence interval) | 2·57 (2·70-4·73) | 2·91 (2·43-3·48) | 2·92 (2·50-3·40) |
| NLR (95% confidence interval) | 0·16 (0·05-0·46) | 0·38 (0·28-0·52) | 0·34 (0·25-0·46) |
| Accuracy | 76·8 (70·7-82·1) | 75·0 (71·9-77·9) | 74·4 (71·6-77·1) |
| **Progressing WHO score** |  |  |  |
| N° events/at risk (%) | 50/228 (21·9) | 221/784 (28·3) | 271/1012 (26·8) |
| Continuously distributed COV50 |  |  |  |
| AUC (95% confidence interval) | 0·76 (0·69-0·83) | 0·76 (0·69-0·83) | 0·72 (0·68-0·75) |
| Categorised COV50 |  |  |  |
| Youden cut-off threshold | 0·04 | 0·04 | 0·04 |
| Sensitivity (95% confidence interval) | 80·0 (66·3-90·0) | 64·7 (58·0-71·0) | 67·5 (61·6-73·1) |
| Specificity (95% confidence interval) | 64·6 (57·1-71·6) | 68·2 (64·2-72·0) | 67·3 (63·8-70·7) |
| PLR (95% confidence interval) | 2·26 (1·77-2·88) | 2·04 (1·74-2·38) | 2·07 (1·81-2·36) |
| NLR (95% confidence interval) | 0·31 (0·18-0·54) | 0·52 (0·43-0·62) | 0·48 (0·40-0·58) |
| Accuracy | 68·0 (61·5-74·0) | 67·2 (63·8-70·5) | 67·4 (64·1-70·3) |

Initial recruitment lasted from 25 March 2020 until 18 November 2020 and continued recruitment from 30 April 2020 until 14 April 2021. NA indicates not applicable. AUC=area under the curve. PLR is the positive likelihood ratio (true positive rate/false positive rate). NLR is the negative likelihood ratio (false negative rate/true negative rate). Accuracy is the overall probability that a patient is correctly classified.

In the whole study population, the AUC of the continuously distributed COV50 urinary marker was 0·81 (95% confidence interval, 0·77-0·85) for mortality and 0·72 (0·62-0·75) for worsening WHO score (table 3). The death rate among participants with a COV50 level less *vs* equal to or higher than the optimised threshold (0·47) was 30/694 *vs* 89/318 (4·32% [95% confidence interval, 4·17-4·48] *vs* 28·0% [27·4-28·6]; p<0·0001); the disease progression rates analysed similarly per optimised threshold (0·04) were 88/587 *vs* 183/425 (15·0% [14·7-15·3] *vs* 43·1% [42·4-43·7]; p<0.0001). The optimised 0·47 threshold for mortality resulted in 74·8% sensitivity, 74·4% specificity and 74·4% accuracy; for worsening WHO score, the optimised 0·04 threshold generated 67·5% sensitivity, 67·3% specificity, 67·4% accuracy (table 3). For both endpoints, these estimates were consistent in the early and continued recruitment phases (table 3).

The odds ratios and discriminatory performance of single baseline risk factors are summarised by recruitment phase in appendix 1 p 12. In the whole study population, on top of sex, age, body mass index, comorbidities, and the baseline WHO score, COV50 analysed as continuously distributed variable and per threshold significantly improved the AUC (figure 1). For mortality, application of the continuously distributed COV50 marker and the 0·47 COV50 threshold, resulted in an increase of the AUC from 0·835 to 0·854 (p=0·0221) and from 0·835 to 0·853 (p=0·0331); for worsening WHO score, the AUC increased from 0·697 to 0·740 (p=0·0001) for the continuously distributed marker and from 0·697 to 0·730 (p=0·0008) for the 0·04 threshold.

**Figure 1:**
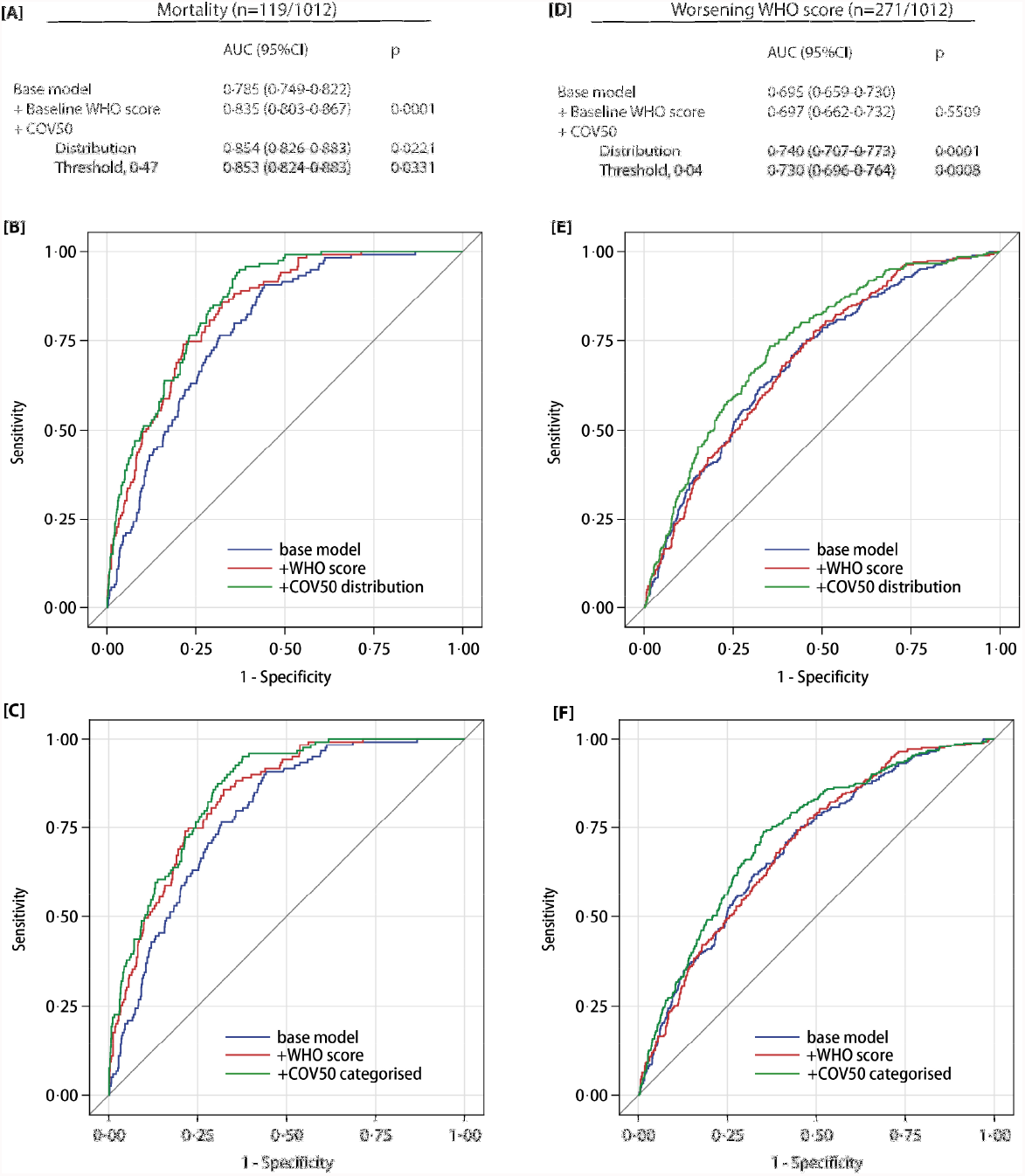
Performance of the COV50 urinary marker on top of other baseline risk factors in the full dataset for contrasting mortality *vs* survival (panels A-C) and for progression *vs* non-progression in the baseline WHO score during follow-up (panels D-F). The base model included sex, age, body mass index and the presence of comorbidities: hypertension, heart failure, diabetes or cancer. In subsequent steps, the baseline WHO score was added and next COV50 as a continuously distributed variable (panels B and E) or as a categorised variable based on an optimised threshold of 0·47 for mortality (panel C) or 0·04 for a worsening WHO score (panel F).

Of 1012 patients, 196 (19·4%) received only ambulatory care, of whom 194 (99·0%) had a baseline COV50 below 0·04; the remaining 816 (80·6%) patients were hospitalised and carried forward in the computation of predicted hospitalisation costs, based on the Markov-chain transition probabilities (appendix 1 p 13) and the simulated number of patients reaching follow-up WHO scores ranging from 3 to 8 (appendix 1 p 14). In low-risk patients (COV50 <0·04), the predicted hospitalisation costs standardised to 1000 patients in regular plus intermediate care compared with intensive care were greater (table 4 and appendix 1 p 15): M€ 6·693 (95% percentile interval, 5·959-7·464) *vs* M€ 5·427 (4·393-6·589). Among high-risk patients (COV50 ≥0·04), the hospitalisation costs showed an opposite structure with lower costs in regular plus intermediate *vs* intensive care: M€ 18·508 (16·952-19·990) *vs* M€ 23·627 (22·196-25·060), in particular in older (≥65 years) high-risk patients (M€ 17·231 [15·625-19·268] *vs* M€ 31·995 [29·562-32·240]).

**Table 4:** Simulated hospitalisation costs by baseline COV50 level, the hospital facility at entry, and age class (starts)

| Baseline COV50<br>Entry hospital facility | Item | Hospital costs expressed per 1000 patients based on the number of patients<br>reaching a follow-up WHO score ranging from 3 to 8 in the Markov chain simulation |  |  |  | Cost reduction<br>associated with 1-day<br>less hospitalisation<br>per 1000 patients |
| --- | --- | --- | --- | --- | --- | --- |
|  |  | 3-4 | 5 | 6-8 | 3-8 |  |
| [-3.26, 3.39[ |  |  |  |  |  |  |
| Regular care | days | 9 (4-45) | 10 (1-37) | 6 (1-51) |  |  |
|  | M€ | 4.304 (4.103-4.504) | 0.906 (0.805-1.006) | 1.956 (1.735-2.189) |  |  |
| Intermediate care | days |  | 6 (3-23) | 11 (1-47) |  |  |
|  | M€ |  | 1.736 (1.543-1.929) | 2.986 (2.649-3.342) |  |  |
| Intensive care | days |  |  | 6 (3-42) |  |  |
|  | M€ |  |  | 9.048 (8.025-10.125) |  |  |
| All care facilities | days |  |  |  | 10 (1-35) |  |
|  | M€ |  |  |  | 20.935 (18.860-23.095) | 2.094 (1.866-2.309) |
| <0.04 |  |  |  |  |  |  |
| Regular care | days | 9 (3-21) | 8 (3-45) | 12 (1-51) |  |  |
|  | M€ | 3.121 (2.946-3.297) | 0.589 (0.514-0.664) | 0.880 (0.712-1.069) |  |  |
| Intermediate care | days |  | 9 (4-19) | 18 (14-22) |  |  |
|  | M€ |  | 1.341 (1.170-1.511) | 0.762 (0.617-0.926) |  |  |
| Intensive care | days |  |  | 4 (3-6) |  |  |
|  | M€ |  |  | 5.427 (4.393-6.589) |  |  |
| All care facilities | days |  |  |  | 10 (3-25) |  |
|  | M€ |  |  |  | 12.120 (10.352-14.053) | 1.212 (1.035-1.405) |
| ≥0.04, [19, 96 y[ |  |  |  |  |  |  |
| Regular care | days | 9 (3-24) | 9 (5-59) | 5 (1-53) |  |  |
|  | M€ | 2.588 (2.300-2.843) | 0.818 (0.658-0.961) | 5.015 (4.711-5.319) |  |  |
| Intermediate care | days |  | 6 (3-23) | 11 (1-47) |  |  |
|  | M€ |  | 1.445 (1.165-1.701) | 8.642 (8.118-9.166) |  |  |
| Intensive care | days |  |  | 6 (3-42) |  |  |
|  | M€ |  |  | 23.627 (22.196-25.060) |  |  |
| All care facilities | days |  |  |  | 11 (3-34) |  |
|  | M€ |  |  |  | 42.135 (39.148-45.050) | 3.830 (3.559-4.095) |

**Table 4: Simulated hospitalisation costs by baseline COV50 level, hospital facility at presentation, and age class (continued)**
| Baseline COV50<br>Entry hospital facility | Item | Hospital costs expressed per 1000 patients based on the number of patients reaching a follow-up WHO score ranging from 3 to 8 in the Markov chain simulation |  |  |  | Cost reduction associated with 1-day less hospitalisation per 1000 patients |
| --- | --- | --- | --- | --- | --- | --- |
|  |  | 3-4 | 5 | 6-8 | 3-8 |  |
| ≥0.04, <65 y |  |  |  |  |  |  |
| Regular care | days | 9 (2-20) | 9 (5-76) | 9 (1-30) |  |  |
|  | M€ | 2.310 (1.897-2.666) | 1.386 (1.134-1.638) | 4.613 (4.135-5.169) |  |  |
| Intermediate care | days |  | 5 (3-17) | 11 (3-25) |  |  |
|  | M€ |  | 2.780 (2.275-3.285) | 6.718 (6.023-7.529) |  |  |
| Intensive care | days |  |  | 5 (3-49) |  |  |
|  | m€ |  |  | 27.582 (24.729-30.911) |  |  |
| All care facilities | days |  |  |  | 12 (3-32) |  |
|  | M€ |  |  |  | 45.389 (40.193-51.198) | 3.782 (3.349-4.266) |
| ≥0.04, ≥65 y |  |  |  |  |  |  |
| Regular care | days | 9 (3-27) | 13 (5-59) | 4 (1-53) |  |  |
|  | m€ | 2.224 (2.214-2.853) | 0.620 (0.421-0.768) | 5.351 (4.944-5.726) |  |  |
| Intermediate care | days |  | 7 (2-24) | 5 (1-47) |  |  |
|  | M€ |  | 1.243 (0.845-1.541) | 7.793 (7.201-8.380) |  |  |
| Intensive care | days |  |  | 12 (1-42) |  |  |
|  | M€ |  |  | 31.995 (29.562-34.240) |  |  |
| All care facilities | days |  |  |  | 11 (2-37) |  |
|  | M€ |  |  |  | 49.226 (45.187-53.508) | 4.503 (4.107-4.864) |

Hospital costs per care facility (median and 95% percentile interval) were extrapolated from the distribution of patients to be expected by Markov chain simulation reaching follow-up WHO scores of 3-4, 5, and 6-8 and the care facility corresponding with disease severity, i.e., regular, intermediate and intensive care for scores 3-4, 5, and 6-8, respectively. Given the simulated number of patients, hospitalisation costs were then computed based on the median number of hospital days per care facility, as observed in the CRIT-Cov-U cohort and a flat facility-specific costing rate harmonised across countries using the 2020 national gross domestic product with the costs in Germany as reference. Hospitalisation costs were standardised to 1000 patients in each risk stratum. Days refers to the median number of days (95% percentile interval) as observed in the CRIT-Cov-U cohort. M€ indicates million Euro.

Measures of treatment efficacy, extracted from the literature review (appendix 2) and exemplary trials (appendix 1 p 16), showed reductions in the number of hospitalisation days or fewer hospitalisation days until recovery. The cost reductions associated with one day less hospitalisation per 1000 patients (table 4) ranged from M€ 1·208 (95% percentile interval, 1·035-1·406) in the lowest risk category (COV50 <0·04) to M€ 4·503 (4·107-4·864) in the highest risk stratum (COV50 ≥0·04 and age ≥65 years).

## Discussion

COV50 is a novel urinary biomarker, consisting of 50 deregulated urinary peptides (appendix 1 pp 8-10). COV50 predicts death and disease progression over and beyond clinical risk factors, comorbidities and the WHO score at presentation. COV50 analysed as single continuously distributed risk factor generated AUCs for mortality and disease progression substantially greater than a 10-year age increment or 1-point increase in the baseline WHO score (appendix 1 p 12). The optimised COV50 thresholds had 74·4% predictive accuracy for mortality and 67·4% for disease progression (table 3). Both COV50 thresholds and COV50 as continuously distributed variable significantly improved the AUC (figure 1).

COV50 is easily administrable, non-invasive, relatively inexpensive balanced against hospitalisation costs, and has high positive and low negative likelihood ratios (table 3). COV50 is registered in Germany and available for clinical and research purposes throughout the European Union. The UPP does not change when urine is stored for 5 days at room temperature in borated test tubes,^9^ thereby providing a wide time window for sample handling, thereby enabling remote testing in non-hospitalised patients with PCR-confirmed COVID-19 infection. UPP profiling requires one day from receipt at the laboratory facility, currently in Hannover, Germany.^9^ Furthermore, for research purposes urine can be indefinitely stored at -20 °C without UPP alteration.^9^ The COV50 test is predictive at the initial stage of infection, when progression to critical illness is still uncertain. Thus, a high-risk test outcome can move patients across the action threshold, allowing the early administration of effective treatments, either in ambulatory or hospitalised care. An extensive literature review (appendix 2) identified effective treatment modalities that might be administered earlier in response to a high-risk UPP risk profile. Such treatments include systemic^10^ or inhaled^11^ corticosteroids, antiviral agents administered as single agents^12^ or cocktails,^13^ monoclonal antibodies antagonising the interleukin-6-receptor^14,15^ or directed against the viral spike protein,^16,17^ or a selective σ1-receptor agonist.^18^ A few exemplary trials were summarised in appendix 1 (p 16).

More than 20,000 peptides detectable in a single urine sample provide a bodywide molecular signature of progressing COVID-19 infection generally independent of the virus strain. A prominent characteristic of the COV50 UPP (appendix 1 pp 8-10) is the shift in collagen fragments, in particular collagen alpha 1(2). Upon infection, the reactive inflammatory cascade activates fibroblasts,^19^ leading to excessive extracellular matrix deposition in response to injury.^20^ The COV50 UPP signature also points to enhanced α1-antitrypsin degradation in line with reports showing that α1-antitrypsin deficiency is associated with life-threatening COVID-19.^21^ Another hallmark of the COV50 UPP is the reduction in urinary peptides derived from CD99.^22^ This observation reflects the loss of endothelial integrity, interference with transendothelial migration of monocytes, neutrophils and T-cells,^22^ and damage of the endothelial tight junctions. The resulting exposure of collagen to the circulating blood triggers the thrombotic complications specific for COVID-19.^23^ As also observed in chronic obstructive lung disease,^24^ with increasing COVID-19 severity, the UPP reveals downregulation of the polymeric immunoglobulin receptor,^22^ which is highly expressed in trachea and the lung and mediates IgA transcytosis.^24^

Cost-effectiveness balances health care costs against non-monetary units, such as quality-adjusted life-years (QALYs).^25^ The QALY-based value proposition is well established in the UK, Sweden, the Benelux and some eastern European countries, but healthcare payers in Germany and France prefer assessing changes in clinical outcomes instead.^26^ CRIT-Cov-U was not designed to address health-economic issues. The administration of quality-of-life questionnaires, the instruments to turn QALY’s into metrics, was impossible in an emergency care setting. Ethics approvals allowing access to claims databases were unavailable. A commonly used definition of the value of human life cannot be applied above age 70 and therefore not to COVID-19-related mortality, given the age distribution of patients.^27^ None of the 52 reviewed trials in COVID-19 patients (appendix 2) included a health-economic analysis. However, the simulations in the current article (table 4) provide some information on the balance between the costs of administering the COV50 test (€850 per test; M€ 0·850 per 1000 patients) against potential health care savings associated with earlier intervention, for instance by the reduction in hospitalisation days, reported to be five up to 10 days in three trials.^13,15,28^ As a high UPP risk profile justifies earlier intertervention over later treatment guided by clinical deterioration, presumably applying the test will not impact on drug costs.

Although COVID-19 affects the heart and kidney, COV50 shares only one urinary peptide with HF1, 13 with CKD273 and two with both classifiers (appendix 1 p 21), which are disease-specific classifiers respectively predictive of left ventricle dysfunction^29^ or chronic kidney disease or diabetic nephropathy.^30^ Thus, COV50 is highly specific for the molecular pathogenic mechanisms associated with SARS-CoV-2 infection. In addition, the high consistency in the discriminatory performance of COV50 among patients recruited initially and later (tables 2 and 3) represents a replication within the CRIT-Cov-U cohort.

Nevertheless, the current results must be interpreted within the context of obvious limitations. First, CRIT-CoV-U is an observational cohort study. Randomised clinical trials should consolidate the optimal strategy for applying treatments guided by COV50 risk profiling. Second, as outlined above, future research should address the health-economic implications of the timing and choice of therapeutic interventions in patients with a low *vs* high risk COV50 score. Third, CRIT-Cov-U enrolled adults and predominantly white Europeans. How ethnicity might affect the UPP is currently under investigation in the Urinary Proteomics Combined with Home Blood Pressure Telemonitoring for Health Care Reform trial (NCT04299529).^31^ Finally, although UPP risk profiling provides insights on the ideal timing of intervention, vaccination remains by far the primordial strategy in addressing the COVID-19 pandemic, although vaccination alone connot be sufficient to restore the pre-COVID-19 way of living.^32^

In conclusion, COV50 is the first biomarker predictive of death and disease progression in adult COVID-19 patients. Independent of clinical risk markers, the operational COV50 thresholds have a discriminatory accuracy or around 70%, even in patients with mild disease. A high-risk COV50 test administered within 4 days of a positive PCR-test justifies earlier treatment in patients with mild-to-moderate disease (WHO scores 1-5), in whom clinical risk factors often leave the prognosis uncertain. Another potential application of COV50 is the selection of patients to be enrolled in randomised clinical trials of novel COVID-19 therapies, in which risk is an issue in the choice between ambulatory *vs* hospitalised care or in the treatment modality to be tested.

## Data Availability

The study protocol is available at the German Register for Clinical Studies (www.drks.de), number DRKS00022495. Anonymised participants data will be made available upon request directed to the corresponding author. Proposals will be reviewed and approved by the funder, investigators and collaborators based on scientific merit. After approval of a proposal, data can be shared through a secure online platform after signing a data access and confidentiality agreement. Data will be made available for a minimum of 3 years after a request has been received and approved.

## Contributors

RW, HM, JM and JB conceptualised the study. HM, JS, JR and JM did the proteomic urine analyses. JAS and LT did the statistical analysis. JAS reviewed the literature and drafted the research-in-context section. JAS, HM and JB wrote the first draft of the manuscript. YLY did the graphics. All authors contributed to the recruitment and follow-up of patients and to the construction and maintenance of the database, and interpreted the results, commented on successive drafts of the manuscript, and approved the final version.

## *Panel:* Research in context

### Evidence before this study

The literature and guidelines were reviewed with as objective to assess the efficacy of interventions in COVID-19 patients relative to the disease stage at presentation. The PubMed search terms ((*COVID-19* OR *SARS-CoV-2*) AND (*clinical* trials OR *randomised trials* OR *randomized trials* OR *RCTs*)) identified 52 articles published in English in 2020 and 2021, which were all read and summarised (appendix 2). Eleven studies enrolled ambulatory patients with mild-to-moderate disease, whereas all other recruited hospitalised patients with moderate-to-severe disease. The median number of days from symptom-onset to intervention varied from 1 to 13 days. Corticosteroids, antiviral drugs, anti-inflammatory and antiviral monoclonal antibodies and fluvoxamine reduced the viral load and disease progression, whereas all other tested drugs and convalescent plasma did not modify the disease course. All studies applied clinical criteria, risk factors, comorbidities or disease-severity scales to stratify for risk. No study implemented a predictive biomarker to triage patients for ambulatory *vs* hospital care or to assess the need of early intervention. Among the directives for the management of COVID-19 patients, only the German guideline mentioned the use of COV50, a urinary proteomic profile biomarker.

### Added value of this study

This study is the first to include a COVID-19-specific biomarker to guide early intervention. COV50 consists of 50 differentially regulated urinary peptides and predicts death and disease progression in adults with mild-to-moderate PCR-confirmed COVID-19 infection (WHO scores 1-5). The predictive accuracy of the optimised COV50 thresholds was 74·4% for mortality and 67·4% for disease progression. On top of covariables and the baseline WHO score, the continuously distributed urinary marker and its optimised thresholds improved the AUCs from 0·835 to 0·854 and to 0·853 for death and from 0·697 to 0·740 and to 0·730 for disease progression, respectively. Using the 0·04 threshold to differentiate low from high COVID-19-associated risk would allow selecting patients with mild disease at presentation for earlier drug treatment, thereby decreasing the risk of worsening disease and death and reducing hospitalisation costs.

### Implications of all the available evidence

The critical question emerging from the 2-year COVID-19 pandemic and most recently from the Omicron variant becoming dominant with high transmissibility is how to prevent deterioration to critical illness in infected patients. A COV50 level of 0·04 or higher predicts disease progression on top of clinical criteria. Even in patients with mild-to-moderate disease (WHO stages 1-5), a high-risk COV50 level is an indication for early in-hospital treatment, thereby valorising the results of the 2020-2021 trials and reducing the burden on health care. COV50 testing can also be applied for the selection of patients in randomised clinical trials of innovative COVID-19 treatment modalities, in which risk at presentation is an issue in the choice between ambulatory *vs* hospitalised care or in the treatment modality to be tested. COV50 is licensed in Germany and available for clinical use in the European Union.

